# Using Deep Learning to Determine Amyloid Deposition through PET and Clinical Data for Alzheimer’s Prognosis

**DOI:** 10.1101/2022.10.04.22280712

**Authors:** Sucheer Maddury, Krish Desai

## Abstract

Amyloid deposition is a vital biomarker in the process of Alzheimer’s diagnosis. Florbetapir PET scans can provide valuable imaging data to determine cortical amyloid quantities. However the process is labor and doctor intensive, requiring extremely specialized education and resources that may not be accessible to everyone, making the amyloid calculation process inefficient.

Deep learning is a rising tool in Alzheimer’s research which could be used to determine amyloid deposition. Using data from the Alzheimer’s Disease Neuroimaging Initiative, we identified 2980 patients with PET imaging, clinical, and genetic data. We tested various ResNet and EfficientNet convolutional neural networks and later combined them with Gradient Boosting Decision Tree algorithms to predict standardized uptake value ratio (SUVR) of amyloid in each patient session. We tried several configurations to find the best model tuning for regression-to-SUVR.

We found that the EfficientNetV2-Small architecture combined with a grid search-tuned Gradient Boosting Decision Tree with 3 axial input slices and clinical and genetic data achieved the lowest loss. Using the mean-absolute-error metric, the loss converged to an MAE of 0.0466, equating to 96.11% accuracy across the 596 patient test set.

We showed that this method is more consistent and accessible in comparison to human readers from previous studies, with lower margins of error and substantially faster calculation times. Deep learning algorithms could be used in hospitals and clinics with resource limitations for amyloid deposition, and shows promise for more imaging tasks as well.

## Introduction

Alzheimer’s disease is a worldwide health concern which has many neurological effects. This common neurological disorder results in brain atrophy, causing patients to experience cognitive decline, behavioral change, and memory loss (Lane & Schott et al. 2018). Diagnosis (particularly early diagnosis) for Alzheimer’s is imperative in order to implement proper treatment plans and delay the progression of the disease (Rasmussen et al. 2019). Efficient and accurate diagnosis is also important in order to save time and reduce error. There is also an overlap in what doctors consider abnormal change and normal age-related change (Mayo Clinic Staff 2022); this creates assessment variability which is an inconsistent practice.

Imaging, clinical data, and physiologic biomarkers are major factors in AD prognosis. Positron Emission Tomography (PET) scans indicate the location of biomarkers in the cerebral cortex. The radiopharmaceutical Florbetapir (18F-AV-45) traces amyloid deposition, an important biomarker which correlates to the progression of Alzheimer’s disease (King Robinson & Wilson et al. 2021).

The Standard Value Uptake Ratio (SUVR) is commonly used as a quantitative measurement of the radiotracer uptake in the brain (Vemuri & Lowe et al. 2016). Pre-existing SUVR values calculated from Florbetapir PET imaging scans were used in our model. To streamline the process of calculating SUVR, we used novel deep learning architecture, a powerful tool that improves efficiency and accuracy in Alzheimer’s prognosis (Saleem & Zahra et al. 2022). We combined deep learning architecture, which was optimized for linear regression, and gradient boosted decision trees to create a SUVR prediction model for this analysis.

## Background Literature

Extracellular amyloid plaques are important in AD characterization (Bloom 2014). Amyloid-β (Aβ) peptides, derived from the amyloid beta precursor protein, are made from amyloid plaques. The accumulation of amyloid plaques disrupt the synapses that facilitate cognition and memory, which show a correlation to similar pathological deficits in AD subjects.

Several studies have examined the relationship between amyloid and AD pathology. Biomarkers use parameters to measure the presence of a disease in a patient. Camus et al. (2012) determined that Florbetapir (18FAV-45) is a core radiotracer biomarker for AD which binds to amyloid plaques. This study found that the mean quantity values of SUVR were higher in AD subjects than HC (Healthy Controls) subjects in cortical regions when using Florbetapir. Because 18F-labeled tracers bind to amyloid plaques, the higher cortical uptake of Florbetapir in MCI and AD subjects compared to HC subjects show that there is a strong correlation between amyloid and AD pathology.

The standardized uptake value ratio (SUVR) is a common way to quantify the severity of a disease. Vemuri et al. (2016) wrote that SUVR is a quantitative measurement which is calculated by the uptake of a radiotracer with respect to the reference region. SUVR can be measured with the uptake values of the Florbetapir 18F radiotracer. Kinahan & Fletcher (2011) quantified the radioactivity concentration from the radiotracer in the region of interest (ROI) over the injection dose of the 18F tracer divided by the weight of the patient with respect to the reference region to measure the SUVR value.

Although studies indicate that an accumulation of amyloid-β corresponds to the characteristics of AD pathology, Ingeno et al. (2019) showed that the removal of amyloid from the brain resulted in the same, or worsened cognitive state when performing clinical trials. However, data on amyloid-β can be utilized for AD prognosis in a given subject.

## Materials and Methods

### General Subject Data

The Alzheimer’s Disease Neuroimaging Initiative (ADNI) provided the Positron Emission Tomography (PET) scans; Mini-Mental State Exam (MMSE) scores; Functional Activities Questionnaire (FAQ) scores; Apolipoprotein (APOE) indication; age, gender, and weight classification that were used in this analysis. ADNI provides biomarker, imaging, clinical, and genetic data across three different groups: CN, MCI, AD. PET scans, MMSE scores, APOE gene indication, FAQ scores, age, gender, and weight were collected for 1298 individuals and 2980 total scans across the amyloid cohort. There were subjects in this cohort that took at least one PET scan. Out of the 1298 individuals from the amyloid cohort, 574 individuals were females and 646 were males.

### Imaging information and SUVR Acquisition

The subjects in the amyloid cohort had the Florbetapir (18F-AV-45) injection for a PET protocol: 370 MBq (10.0 mCi) ± 10%, 20 min (4×5min frames) acquisition at 50-70 min post-injection.

For each subject, all scans were collected from ADNI’s image and data archive using a specific advanced search (“AV45 Coreg, Avg, Std Img and Vox Siz, Uniform Resolution“). The scans from this search were coregistered PET-MR and intensity normalized images that used Statistical Parametric Mapping (SPM8), a process which performs many voxel-wise comparisons to assess the significance of cerebral blood flow changes (Vieira et al. 2020). Over the 20 minute acquisition time, each image was resized to a uniform voxel size and each uniform size was 160×160 in-plane, along with 96 axial slices (Landau et al. 2021; Reith et al. 2020). All images were normalized and rescaled to 224×224 to accommodate the ImageNet pretraining.

We obtained the Florbetapir cortical summary SUVR (“SUMMARYSUVR_WHOLECEREBNORM”) for each scan from the UC Berkeley AV45 Analysis. This calculation required FreeSurfer processing which included skull-stripping, segmentation, and delineation of cortical and subcortical regions in MRI scans which were coregistered to PET scans using SPM8. The cortical summary region (“COMPOSITE_SUVR”) was calculated by taking the mean uptake of all SUVR values from the subregions. These SUVR (“COMPOSITE_SUVR”) values were calculated with respect to the reference region (“WHOLECEREBELLUM_SUVR”) to derive the summary SUVR value for the whole cerebellum (“SUMMARYSUVR_WHOLECEREBNORM”) for each scan (Landau et al. 2021).

**Equation 1:** Standardized Uptake Value Ratio.

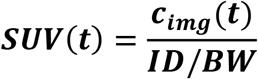

SUV(t) represents the radioactivity concentration in a region during the period of time over the quantity of the injected dose (kBq/mL) divided by the weight (kg). This ratio is calculated with respect to the reference region. The SUVR is visualized below in **Figure 1**.

**Figure 1:**
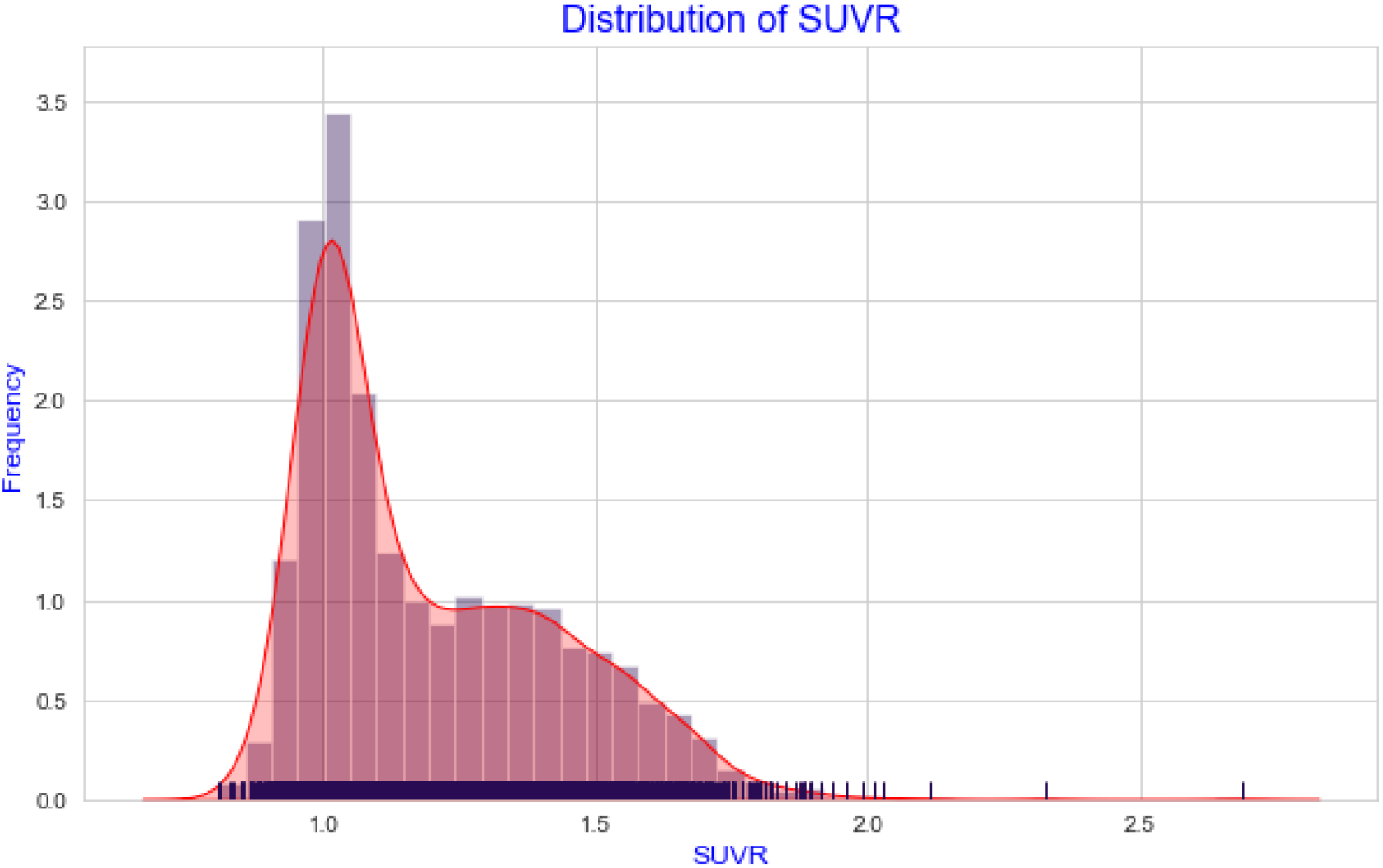
Distribution of the SUVR values.

### Clinical Data

An individual’s age, gender, and weight were included in the clinical data for this analysis. Each individual in the ADNI dataset received a Mini-Mental State Exam (MMSE) after their testing session. CN or MCI subjects normally score between 24 and 30 inclusive while AD subjects normally score between 20 and 26 inclusive, showing that subjects who score lower than normal on this exam have cognitive impairment which is an indicator of Alzheimer’s (Peterson et al. 2010).

Individuals also took a Functional Activities Questionnaire (FAQ) after their testing session. FAQ tests subjects with daily activities; the questionnaire has a range of 0-30 and subjects with a score of 6 or greater is suggestive of functional, cognitive impairment (Marshall et al. 2015).

Apolipoprotein E is a multifunctional protein with three isoforms: APOE ε2, APOE ε3, and APOE ε4. APOE ε4 has the possibility of forming stable complexes with Aβ peptides and it enhances Aβ aggregation. This suggests that there is a correlation between APOE ε4 and pathogenesis of AD (Huang et al. 2014). While APOE ε4 is more of a genetic risk factor AD, subjects with APOE ε3 are generally neutral and subjects with APOE ε2 are protective (Huang et al. 2017).

### Deep Learning Implementation

The deep learning was implemented using TensorFlow (https://www.tensorflow.org/). The data was split into training (80%, n=2384), and testing (20%, n=596) subsets in order to isolate training and testing results. We first used the ResNet convolutional neural network (CNN) architecture, which solves the vanishing/exploding gradient problem via skip connections (He, Zhang, Ren, & Sun 2015). Skip connections calculate the identity function of an earlier layer output and add it to the output value of the succeeding layer, preserving the gradient (Adaloglou 2020). Skip connections (per block) are represented by the following function:

**Equation 2:** RELU activation.

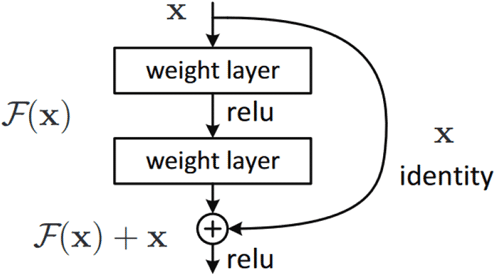

We also tested EfficientNet, which uses compound model scaling to achieve better results than ResNet (Tan & Le 2021). EfficientNet uses a specific set of scaling coefficients to uniformly scale the resolution, width, and depth in order to achieve a constant ratio (Sarkar 2021).

**Equation 3:** Compound model scaling.

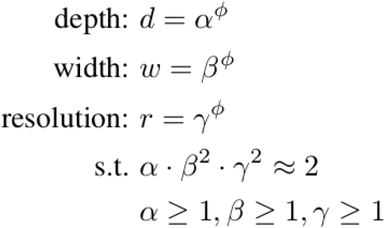

^1^ Conditions for **α, β**, and **γ** where Φ is a user-specified coefficient

Adam was used to optimize loss via backpropagation (Kingma, & Ba 2014). An initial learning rate of either 0.001 or 0.0003 was used with a batch size of 32. All models were pre trained on ImageNet weights.

### Gradient Boosted Decision Trees

Gradient Boosted Decision Trees sequentially build simple prediction models while constantly correcting the preceding model. This process improves the mistakes of the previous learner while simultaneously filtering out the correct observations (Gaurav 2021). LightGBM is an open-source library that provides automatic feature selection and larger gradients which improves predictive performance of gradient boosted decision trees (Brownlee 2020).

The GBM (Gradient Boosting Machine) was trained for 50,000 iterations with an early stopping sensitivity of 500 iterations. A random grid search was used to find the optimal hyperparameters for the GBM, by substituting random parameters and evaluating which parameters performed the best. Random state variables were never tested, with the intent to preserve scientific integrity.

### Prediction Approaches

Several prediction approaches were used with the data. First, ResNet50 was used to classify amyloid positivity in a single slice. Slice 48 was chosen out of the 96 axial slices, as it covers the central region of the brain which Alzheimer’s often affects. The proposed cutoff value of 1.1 for SUMMARYSUVR_WHOLECEREBNORM was used (Landau & Jagust 2015). The average pooling layer preceding the fully connected layer outputs 2048 activations in the standard ResNet model. The fully connected layer was changed to down sample the 2048 activations to 2 classes through linear down sampling. Binary CrossEntropy was used as the loss function:

**Equation 4:** Binary Cross Entropy Loss.

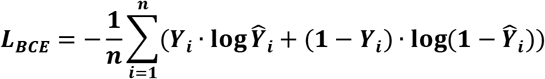

Second, regression to SUVR was performed with three slices, slice 36, 48, and 60. Color composites were created by overlapping slices 36, 48, and 60 into the R, G, and B color channels respectively. Since the images are all black and white (thus governed by one color channel), no imaging information was lost by doing this, and during the prediction, the model will split the image into their respective color channels regardless, effectively providing three images worth of information in one. Examples are shown in **Figure 2**.

**Figure 2:**
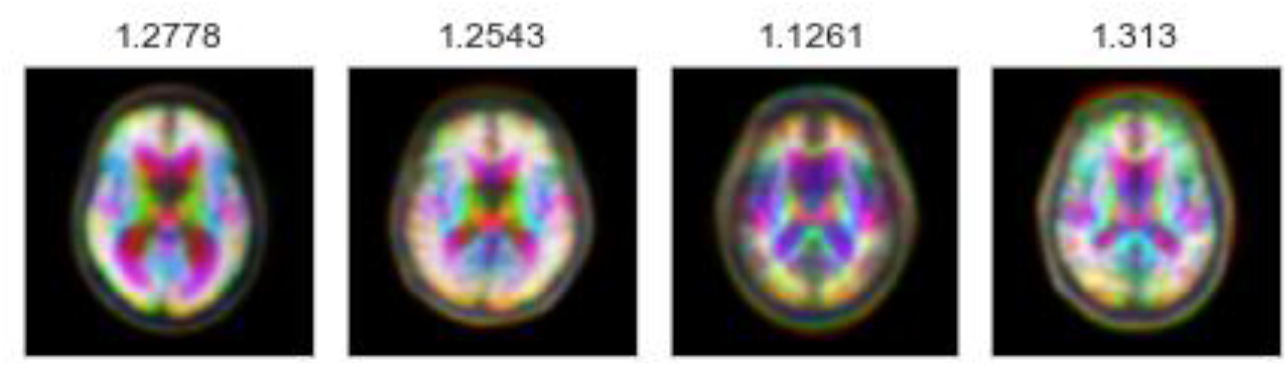
Color composites of various subjects with ground truth SUVR values.

For regression, the last fully connected was changed to one output which is linear only. Mean Absolute Error (MAE) was used to measure regression loss. MAE is the average difference between predicted and ground truth values, used in order to quantify the average difference between a patient’s true SUVR value in the field versus the model prediction.

**Equation 5:** Mean Absolute Error.

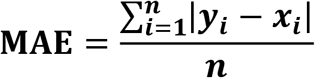

Finally, the best performing architecture was once again trained on the RGB color composites. The last fully connected layer was then removed and the activations were extracted from the GlobalAveragePooling layer. The activations, as well as the clinical and genetic data, were fed into the Gradient Boosted Decision Tree, which then performed regression to reach a SUVR value. In effect, the linear layer was being replaced by GBDT functions, which has been shown to be more accurate (Ke et al. 2017).

## Results

### Binary Classification

First, we trained on binarized amyloid classification for SUVR (positive/negative), found using the cutoff value discussed above. Binary classification can be useful in determining positivity of Alzheimer’s, although it lacks to precision of an exact SUVR value. Using pre-trained ImageNet weights, 30 epochs, and a batch size of 32, we used an initial learning rate of 0.001 to achieve an accuracy of 100% for the training set and an accuracy of 91.8% for the testing set. The CrossEntropy losses for training and testing set after 30 epochs were 0.0 and 0.444, respectively. Out of 596 subjects from the testing set, 547 subjects were classified correctly. Accuracy is plotted in **Figure 3**, which seems to have leveled off for the training and testing sets after 15 epochs, likely because of the simplistic binary task.

**Figure 3:**
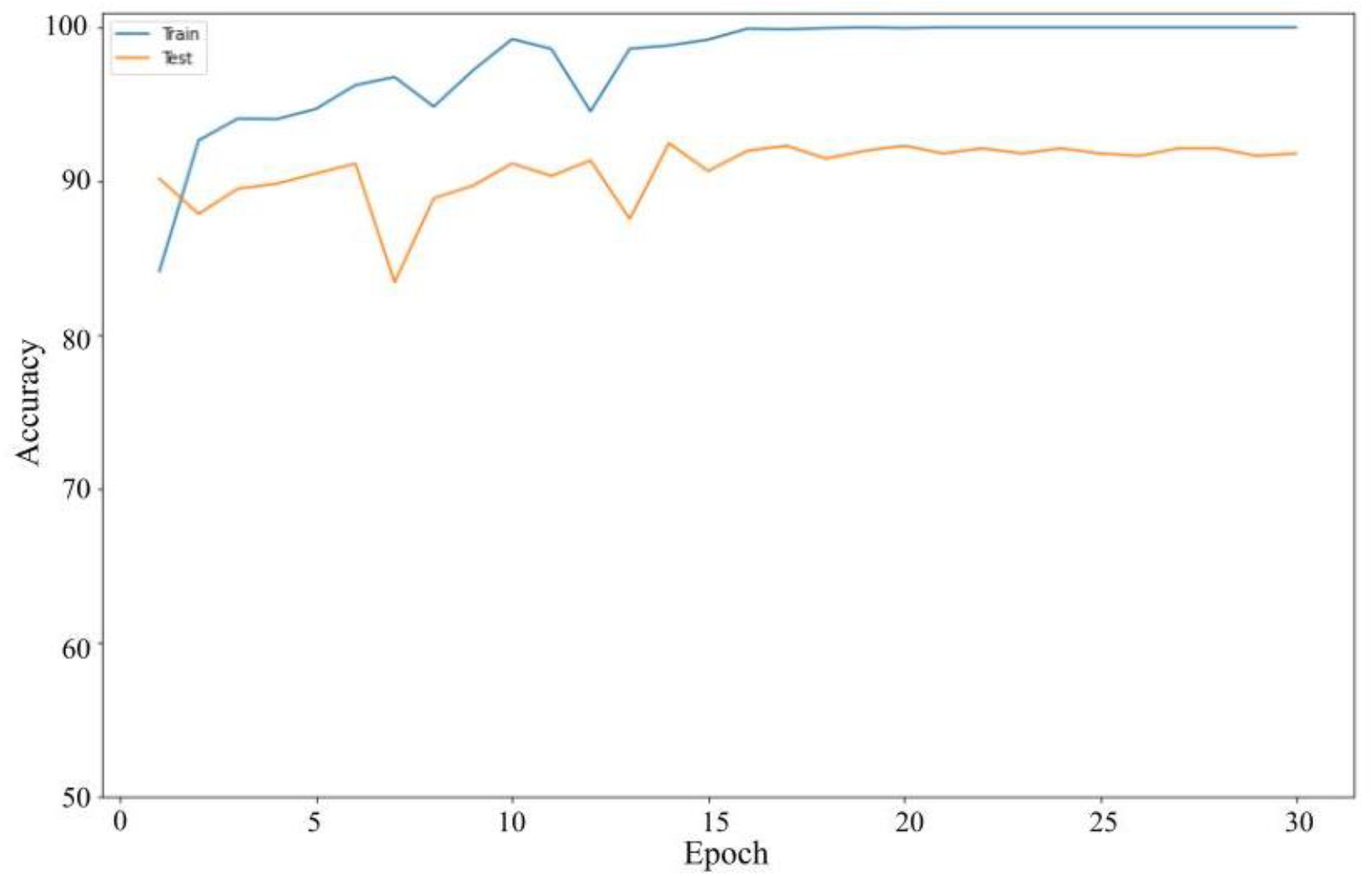
Binary Classification Accuracy for Amyloid Positivity/Negativity up to 30 epochs

### Amyloid Regression Model

For regression we used 100 epochs, and an initial learning rate of 0.0003. We used four different regression models: ResNet50, ResNet101, ResNetRS50, and EfficientNetV2B3. ResNetRS is a modern revision of the original ResNet architecture, outperforming the original ResNet by ~4.5% (Bello et al. 2021). EfficientNetV2 is the updated version of the original EfficientNet Architecture, using less memory and achieving better and faster results. The MAE loss for the training set and testing set of the four regression models are shown in **Table 1**.

**Table 1:** Various Linear Regression Model MAE Loss Results for training and testing sets

| CNNs | MAE for train set | MAE for test set |
| --- | --- | --- |
| **ResNet50** | 0.0380 | 0.0665 |
| **ResNet101** | 0.0341 | 0.0580 |
| **ResNetRS50** | 0.0320 | 0.0533 |
| **EfficientNetV2B3** | 0.0312 | 0.0517 |

<sup>1</sup> Each trial was run a single time and results were taken (n=2980).

The best results were achieved on the EfficientNetV2 architecture. EfficientNetV2 was the smallest of all the models with only ~10 million parameters, yet outperformed substantially compared to more costly models, such as ResNet101 and ResNetRS50. The increase in layers from 50 to 101 in ResNet decreased error, although outperformed by the more costly ResNetRS architecture. This seems to suggest that the compound scaling mechanism of the EfficientNet architecture is effective at scaling layers based on the image size. Additionally, the difference between the train set and test set accuracy was lowest in the EfficientNetV2B3 model, signifying less overfitting.

Minimal consistency improvements were made after 60 epochs, thus we decided to train for 60 epochs in succeeding tests. The EfficientNetV2 architecture performed the best out of all the models. **Table 1** shows that EfficientNetV2B3 achieved the lowest MAE loss value compared to the other three regression models. EfficientNetV2B3 had a 3% improvement compared to the best residual neural network. This improvement is also visually shown in the epoch vs loss graphs for training and testing sets with the four regression models in **Figure 4**. Because of this, EfficientNetV2B3 was the architecture that was used with the Gradient Boosted Decision Tree.

**Figure 4:**
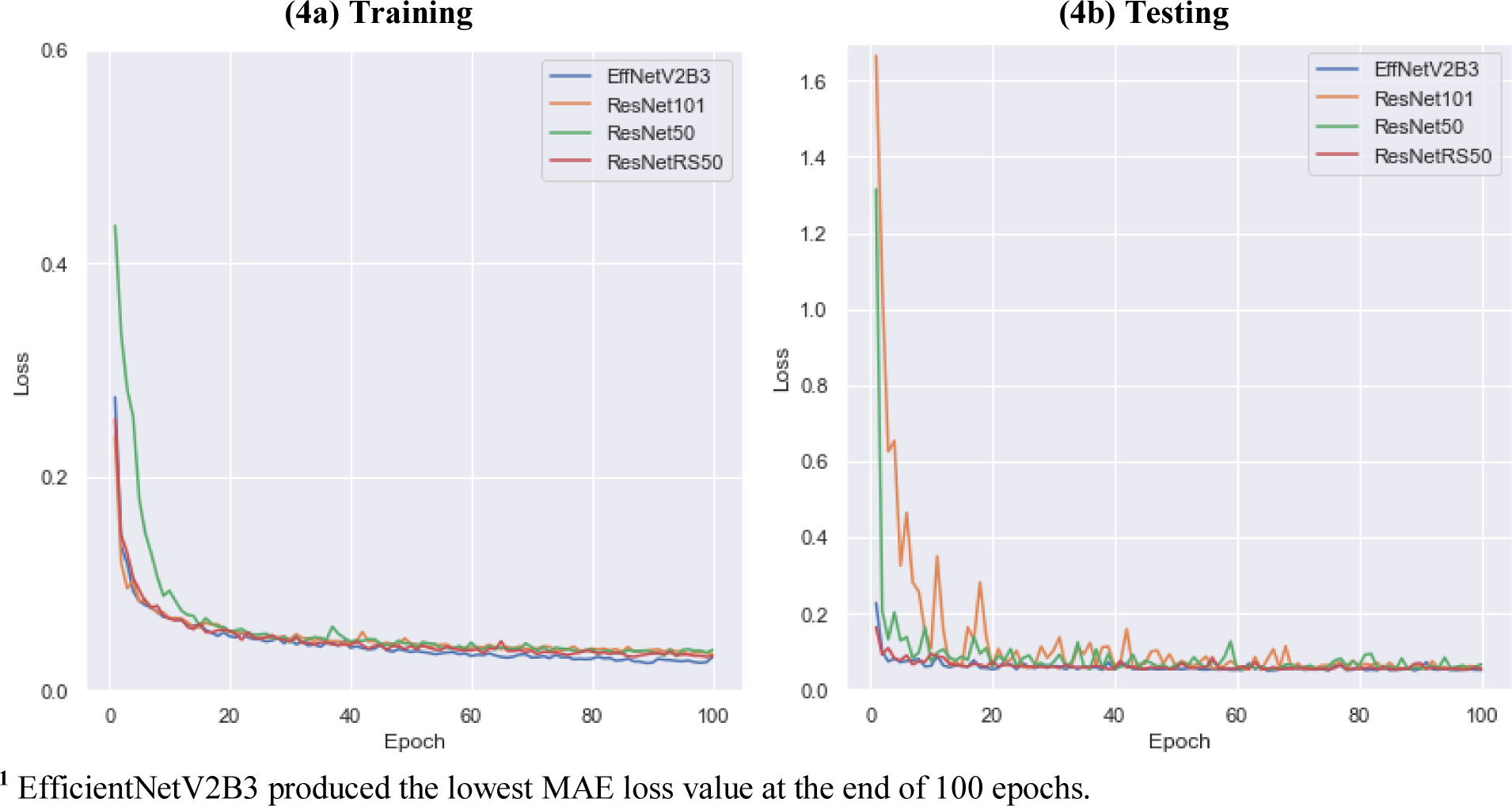
Epoch vs loss for training and testing sets of ResNet50, ResNet101, ResNetRS50, and EfficientNetV2B3.

**Figure 5:**
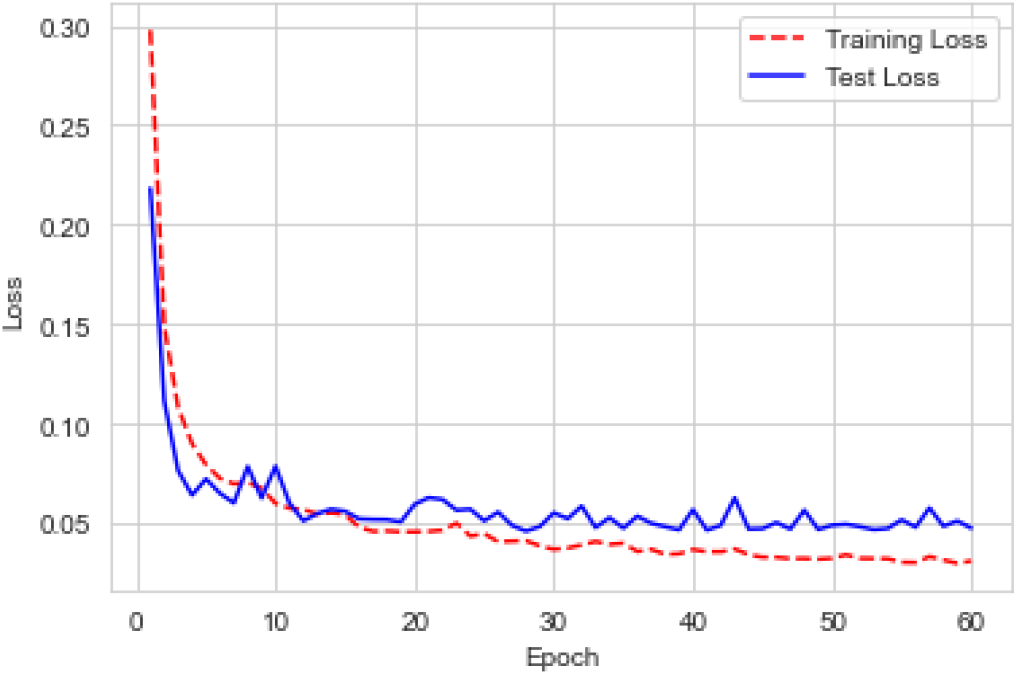
Epoch vs loss for training and testing sets of the EfficientNetV2S CNN

### Amyloid Regression and Gradient Boosted Decision Tree Model

We used the EfficientV2S network for regression and gradient boosted decision trees for this model. The EfficientV2S regression model used 60 epochs and an initial learning rate of 0.0003. GlobalAveragePooling was the layer preceding the fully connected layer, outputting 1280 activations per subject in the EfficientNetV2 architecture. This regression model achieved an MAE loss value of 0.0477 for the testing set, already a significant improvement from all previous models. Model accuracy improved insignificantly after 30 epochs but did have noticeable consistency improvements.

We then used the LightGBM library for our GBDT because it has improved predictive performance compared to other GBDTs. We used clinical data with LightGBM. We also used random grid search for 100 iterations to find the best hyperparameters for the GBDT model which resulted in the lowest MAE loss. The optimal hyperparameters were a max_depth of 9, a feature_fraction of 0.5, and a learning_rate of 0.045. With EfficientV2S and LightGBM, the MAE loss value for the testing set was 0.0466, outperforming all previous configurations by a high margin and improving the loss from the CNN by 0.0011. The basic model flow is shown below in **Figure 6**.

**Figure 6:**
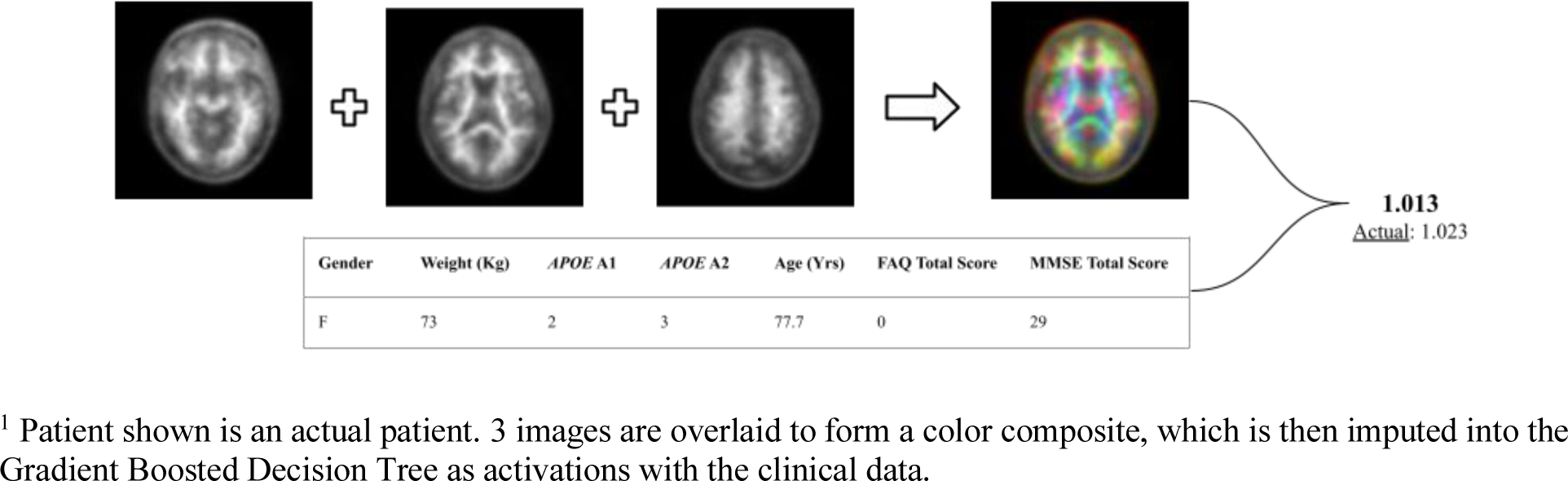
Basic model diagram.

To observe the consistency improvements in **Figure 6**, a predicted vs ground truth plot was drawn using all 2980 data points. This way, any outliers could be spotted and skews could be identified. The model shows a consistent, fairly tight correlation between the ground truth and predicted values, as shown in **Figure 7**, with little to no skew.

**Figure 7:**
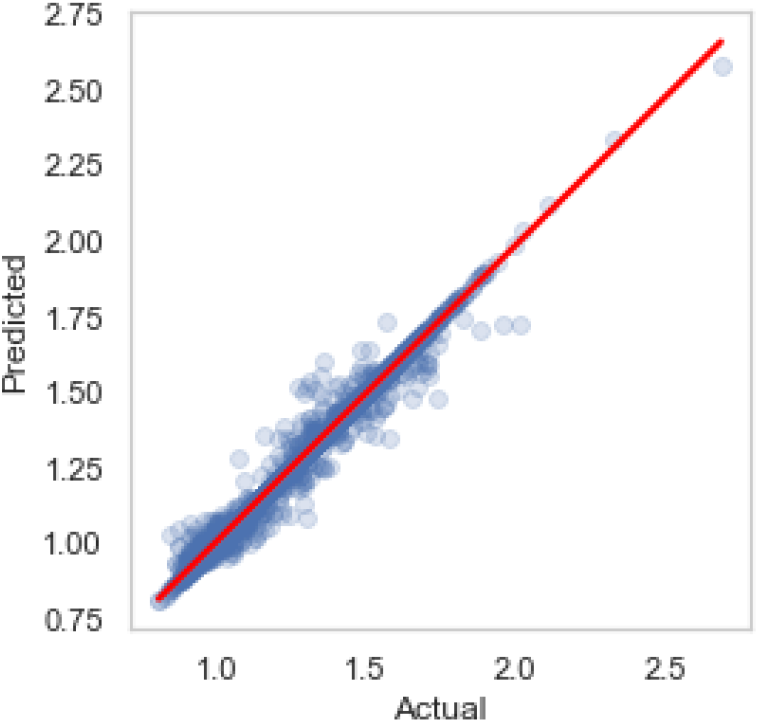
Predicted vs Actual for Efficient V2S and LightGBM model.

## Discussions

Optimizing a deep learning network for linear regression is a more efficient and accurate way to predict SUVR. Reith et al. (2020) used a convolutional neural network (ResNet-50) to predict SUVR by optimizing network depth, using 3 axial slices per subject and ImageNet pretraining. This model achieved an RMSE loss value of 0.054 for the SUVR prediction.

We first used four regression models (ResNet50, ResNet101, ResNetRS50, and EfficientNetV2B3) to see which network architecture is best for MAE loss. Similarly to Reith et al. (2020), we used 3 axial slices per subject and ImageNet pretraining. However, our slice selection was different from Reith et al. (2020). We used axial slices 36, 48, and 60; the areas of these slices have the highest amyloid burden in subjects. We found that EfficientNetV2B3 performed the best out of all four models with an MAE loss of 0.0517 for the testing set.

After determining that EfficientNetV2 is the best network architecture, we used the EfficientNetV2S network which has twice as many parameters than EfficientNetV2B3. We used axial slices 36, 48, 60 and ImageNet pretraining for this network and achieved an MAE loss of 0.0477. EfficientNetV2S had a 17.8% improvement compared to the most accurate residual neural network and a 7.7% improvement compared to EfficientNetV2B3.

Consequently, we used the LightGBM library for GBDT after determining that EfficientNetV2S is the highest performing convolutional neural network. For LightGBM, we implemented a random grid search algorithm which found the optimal hyperparameters for GBDT. We also added three new variables: MMSE scores, FAQ scores, and APOE indication. This resulted in an MAE loss of 0.0466 for the testing set which outperformed our best CNN model by 0.0011.

Results from our study show that the use of axial slices 36, 48, 60 per subject, MMSE scores, FAQ scores, APOE indication, and LightGBM paired with EfficientNetV2S improved the linear regression model’s prediction performance of SUVR significantly. Our best regression model (0.0466 MAE loss) achieved an accuracy of 96.1%. Our proposed regression model (EfficientV2S and LightGBM) improved by 22.3% compared to the network in the Kim et al. (2019) study and improved by 13.7% compared to the Reith et al. (2020) study.

Although the calculation of SUVR for a given subject provides the uptake quantification of the radiotracer (18F-AV-45) in malignant cells, this calculation approach is inefficient and less accurate compared to a deep learning approach. When comparing SUVR prediction performance from a linear regression model to SUVR calculations by readers, Reith et al. (2020) found that the three SUVR readers took 24:28 minutes for 100 test samples. Our study used 600 test samples and took ~18 seconds while the SUVR readers would have taken ~147 minutes to calculate all SUVR values. Individual SUVR calculations are not ideal when diagnosing a patient with a Florbetapir PET scan. Our proposed model solves the efficiency problem that SUVR readers experience when calculating SUVR values.

Noise in the ground truth SUVR calculations for each subject’s scan needs to be considered with the result of the regression model. Reith et al. (2020) showed that each reader calculated the SUVR value at a different pace and accuracy which contributes to the SUVR variability factor. There was also noticeable noise in the Florbetapir PET scans. The pixel count was 160×160 which is not a very clear resolution. The scans were also in black and white, another factor which might have contributed to the noise of the scans. There was noise in the chosen slices because there might have not been enough coverage for parts of the brain which have more present amyloid or are highly correlated to AD.

There are several limitations to consider in this study. Firstly, we were only able to examine the information in the input and the output layers of the CNN but not the middle layers which are responsible for tasks such as data transformation and automatic feature creation. For future use of this model, images fed as input data would require a specific process. Each Florbetapir image needs to be co-registered using Statistical Parametric Mapping (SPM8) to the same subject’s MRI image. This process requires the subject to get a PET and MRI scan. Also SPM8 software is necessary for the co-registering process. This process alone questions the practicality and financial cost of the imaging (Landau et al. 2021).

## Conclusions

Ultimately, we used deep learning architecture and Gradient Boosting Decision Trees along with imaging, clinical, and SUVR data to construct a regression model. The proposed model predicts SUVR values of subjects at a higher accuracy and efficiency than previous studies. Our model quantifies amyloid from Florbetapir radiotracer uptake in PET scans. Our automated model can overcome the difficulties of quantifying SUVR for patients with a Florbetapir PET scan. Our model can be used for processing large amounts of data from clinical trials. The achieved accuracy from our model provides greater reliability compared to calculations made by trained SUVR readers. Future work includes an application which allows a scan to be fed into the network in order to predict the SUVR quantification with our best performing linear regression model. Future research should investigate a more accurate way to individually calculate SUVR values so that deep learning networks can provide better accuracy for the regression model. Solving noise factors should be considered in future research in order to limit variability in SUVR and make PET visualizations better quality. Better PET imaging would also result in better accuracy from regression models when training the network.

## Data Availability

All data and code is available at:

https://adni.loni.usc.edu/

https://github.com/sumaddury/Amyloid-PET-SUVR-Quantification

## Author Contributions

S.M.: Conceptualization, data collection and processing, machine learning, optimization, visualization, paper editing. K. D.: Conceptualization, data collection, methodology, background research, visualization, validation, paper writing and editing. All authors contributed to and have read and agreed to the final version of the manuscript.

## Acknowledgements

We would like to thank Caitlin Corona-Long (M.A.) for her mentorship and support, and Dr. Arnold Bakker for his consultancy during publication and revision.

